# Dental caries among the elderly in Iran: a meta-analysis

**DOI:** 10.1101/2020.07.24.20161299

**Authors:** Shervan Shoaee, Farshad Sharifi, Pooneh Ghavidel Parsa, Ahmad Sofi-Mahmudi

## Abstract

**Objectives:** To conduct a systematic review and meta-analysis on dental caries among the elderly in Iran.

**Background:** The prevalence of dental caries among elderly is high worldwide, and the major burden of oral diseases is caused by dental caries.

**Methods:** Systematic review of the published and grey literature performed. Six international and local databases used to provide the most comprehensive population-based studies. National oral health surveys, as well as national disease and health surveys considered as other primary sources of data. Quality of remained studies was assessed by a modified tool designed based on STROBE statement checklist to evaluate the cross-sectional studies. The target population was 65+-year-olds Iranian population.

**Results:** Overall, 917 English articles who those reported dental caries among all age groups were found in international databases and 2138 Persian articles were found in Iranian databases. After quality assessment, and excluding other age groups, 46 points of data with 10411 aged people ≥ 65 years, were included in the meta-analysis. Mean pooled decayed, missing, and filled teeth among the elderly was 26.84 (26.41-27.28). This index was 26.78 (26.12-27.43) in women and 26.91 (26.32-27.50) in men. Mean number of decayed teeth was 1.48 (1.32-1.65). Mean pooled missing teeth was 24.83 (24.20-25.46), and mean pooled filled teeth was 0.14 (0.12-0.17). The majority (92%) of DMFT was related to missing teeth.

**Conclusion:** Dental caries has a very high burden on the oral health of the elderly in Iran, mainly responded by tooth extraction.

## Introduction

Dental caries are a global health condition affecting both children and adults which can lead to disability. Despite being largely preventable, the prevalence of dental caries among adults is high worldwide affecting 35% of the global population which makes it the most prevalent health condition in the world (1). According to the World Health Organization (WHO), dental caries among older adults aged 65 years or more remains a significant problem for countries within WHO regions. The mean dental caries experience (Decayed, Missing and Filled Teeth or DMFT index) among this population group across Eastern Mediterranean region (which Iran is located) is reported about 23, and the prevalence rate of edentulism is around 30% (2). According to studies conducted in Iran, the prevalence of edentulism was estimated to be between 60-78% among people over age 70 years (3, 4).

The impact of dental caries on individuals and communities as a result of the pain and suffering, impairment of functions and communication, and reduced quality of life are considerable (5, 6). As well as dental caries, periodontal diseases could lead to edentulism and impairment of social functions (7-9). Furthermore, several oral conditions are linked to non-communicable diseases (NCDs) primarily because of their common risk factors (10). General diseases often have oral manifestations (e.g. diabetes or HIV/AIDS) (11). The relationship between periodontal diseases or tooth loss, and increased risk of cardiovascular diseases (CVD), pulmonary diseases, diabetes, pregnancy outcomes, and all-cause mortality have become a constant issue of discussion in the current scientific literature (12, 13).

Advances in oral health care and treatment in the past few decades have resulted in a reduced number of edentulous individuals and the proportion of adults who retain their natural teeth until late in life has increased substantially (14, 15). Besides, a still growing number of dentate older people have tooth wear, oral implants, sophisticated tooth- and implant-supported restorations and prostheses (16). Hence, they are in continuous need of both preventive and restorative oral health care. The complexity of oral health status, systemic diseases, and the use of multiple medications make elderly people more vulnerable to oral problems than younger age groups, even more in those who are cognitively impaired (17).

The magnitude of disease distribution provides a unique perspective for planning interventions and developing public health policies. One of the most crucial research methods to gain an accurate estimation of indicators (indices) of disease in society is a structured review and meta-analysis. Based on our knowledge, no meta-analyses have been done to combine results of the mean of DMFT among old people in the general population of Iran. The present study aims to conduct a systematic review and meta-analysis to determine the mean of DMFT among elderly Iranian people over 65 years.

## Methods

This research is a sub-component of the National And Sub-national Burden of Oral Diseases (NASBOD) study. In the NASBOD study, a systematic review of published literature was conducted, as well as using unpublished and grey literature. We followed recommendations for reporting of meta-analysis of observational studies in epidemiology (18).

### Search strategy

Three international databases (PubMed, Web of Science, and Scopus) and three national databases (IranMedex, SID, and Irandoc) were used to find studies and provide the most comprehensive epidemiological data bank on 17 March 2019. The search through international databases was performed using MeSH terms, Entry terms, Emtree, and related keywords. Persian keywords were mostly developed based on their English equivalents, and some expert panels were also conducted to review any probable keywords. The main English keywords included “dental caries”, “DMF index”, “dental restoration, permanent”, “tooth diseases”, and “dental health surveys”.

### Selection criteria

The next steps were based on title exclusion, abstract exclusion, and reviewing of full texts. The full texts of all relevant articles were extracted, and a comprehensive quality assessment form was designed to assess the quality of selected articles.

### Study design

Only population-based studies reporting dental caries in Iran were included.

### Participants

The target population in the systematic review was representative Iranian population of any ages, but for the meta-analysis, we entered studies reported dental caries for those over 65 years old. Another primary source of data was the National Oral Health Surveys providing dental caries for the elderly.

### Outcomes

The main outcome of interest was DMFT. As the main component of DMFT in the elderly is missing teeth, many studies had only reported this component. However, we reported all components altogether and separately.

### Selection of searched articles

Following the search in selected search engines, articles were selected according to the inclusion and exclusion criteria in three phases. In case of disagreement, reviewers discuss the indefinite articles to reach an agreement and select them. Full texts were obtained via referring to the digital library of Tehran University of Medical Sciences, contacting with corresponding authors or accessing to the journal in which the article was published.

#### Title phase

Reviewers scanned all titles according to the inclusion criteria. Titles in doubt were also included.

#### Abstract phase

Reviewers scanned all abstracts according to the inclusion criteria. Articles with unclear methods were also included in this phase.

#### Full-text phase

To finalise the selection phase, reviewers evaluated all remaining articles to find out if they were eligible.

### Quality assessment and data extraction

Author, province, year of publication, year of study, duration of study, sex, age, DMFT index, decayed teeth, missing teeth, filling teeth, DMFT index examiner, methods of examining decayed teeth, methods of examining missing teeth, and methods of examining filling teeth were independently extracted from included studies by two of authors. Any disagreement, if were not resolved by consensus, a third author decided about it. Although DMFT index is usually determined by examining 28 teeth in the oral cavity (from central incisor to the second molar in each quadrant), these data have taken into account the third molars so that the highest end of DMFT index in our data is 32 instead of 28.

Quality of included studies was assessed by a form derived from STrengthening the Reporting of OBservational studies in Epidemiology (STROBE) (19). A copy of this form is available in Appendix 1.

### Data synthesis and analysis

We used R version 3.6.0 for Windows with meta (4.9-6) and metafor (2.1-0) packages for statistical analysis. We calculated the mean of each parameter (DMFT and its components).

Heterogeneity was assessed using Cochran’s Q and F statistics (20). For assessing publication bias and selective reporting, we used funnel plots and Egger’s regression intercept test. A p-value below 0.05 was considered as statistically significant when applicable.

## Results

At first, we reached 3055 articles (917 English and 2138 Persian) reporting dental caries among all age. By removing duplicates, 947 articles remained. After quality assessment and excluding other age groups, 46 points of data with aged people ≥ 65 years, including two from cross-sectional studies (conducted in two northern provinces in 2009) and 44 from two national data, one conducted in 13 provinces in 1999 and the other in 30 provinces in 2012, were included in the meta-analysis (Figure 1).

**Figure 1.**
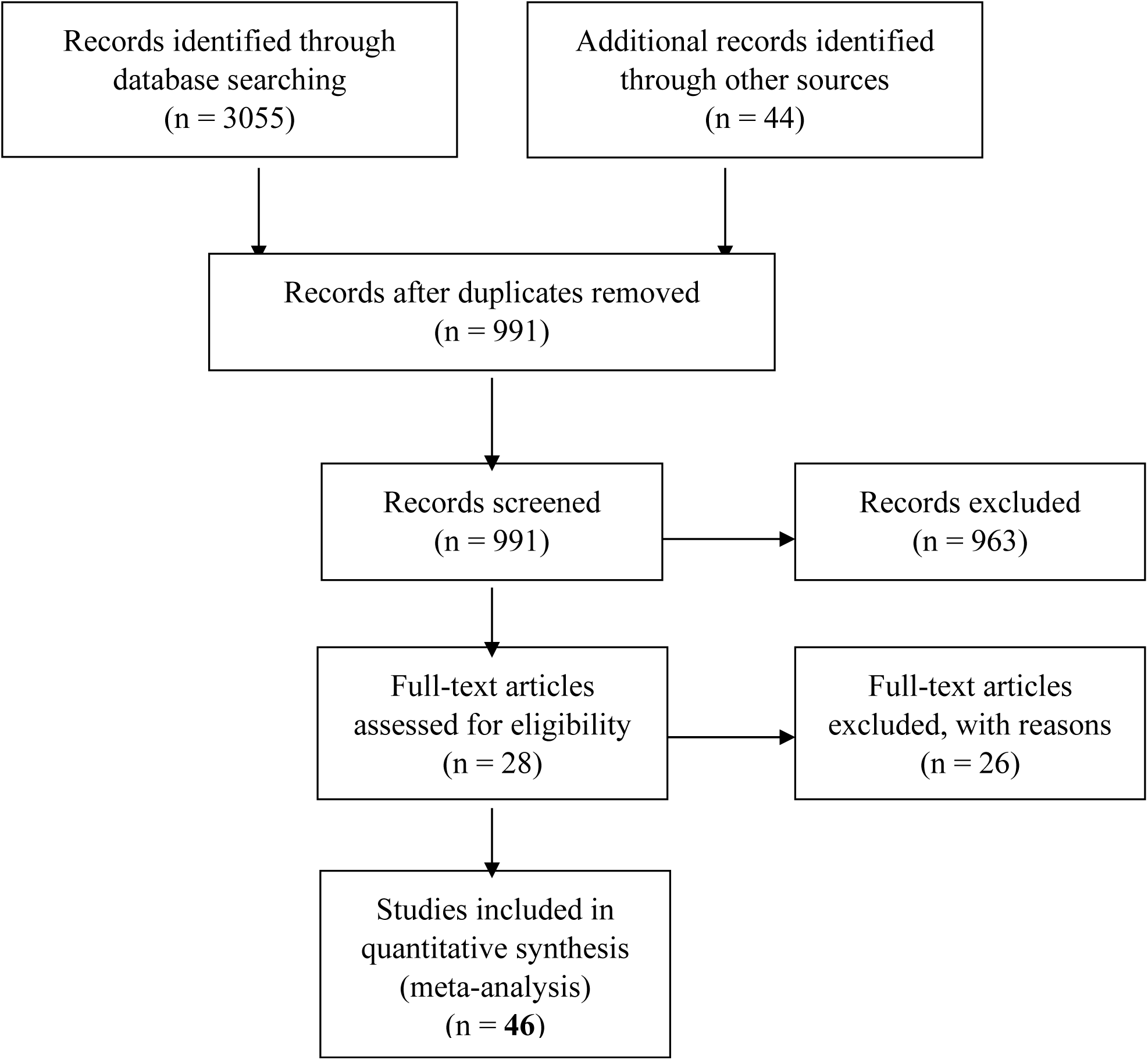
Study selection flowchart.

The total elderly population of this meta-analysis were 10411 (50.67% female). The pooled mean of DMF in the Iranian population at the national level by using random effect method was 26.84 (95% CI 26.41 to 27.28; I^2^ = 87%, n = 9542). The mean DMF according to gender was 26.78 (95% CI 26.12 to 27.43; I^2^ = 88%, n = 4862) for females and 26.91 (95% CI 26.32 to 27.50; I^2^ = 85%, n = 4680) for males. The mean of each component of DMF was as follows: dental decay: 1.48 (95% CI 1.32 to 1.65; I^2^ = 90%, n = 9339); missing teeth: 24.83 (95% CI 24.20 to 25.46; I^2^ = 91%, n = 10411); and filling teeth: 0.14 (95% CI 0.12 to 0.17; I^2^= 77%, n = 9339). The forest plots for DMF and its components by gender are shown in Figures 2 to 5.

**Figure 2.**
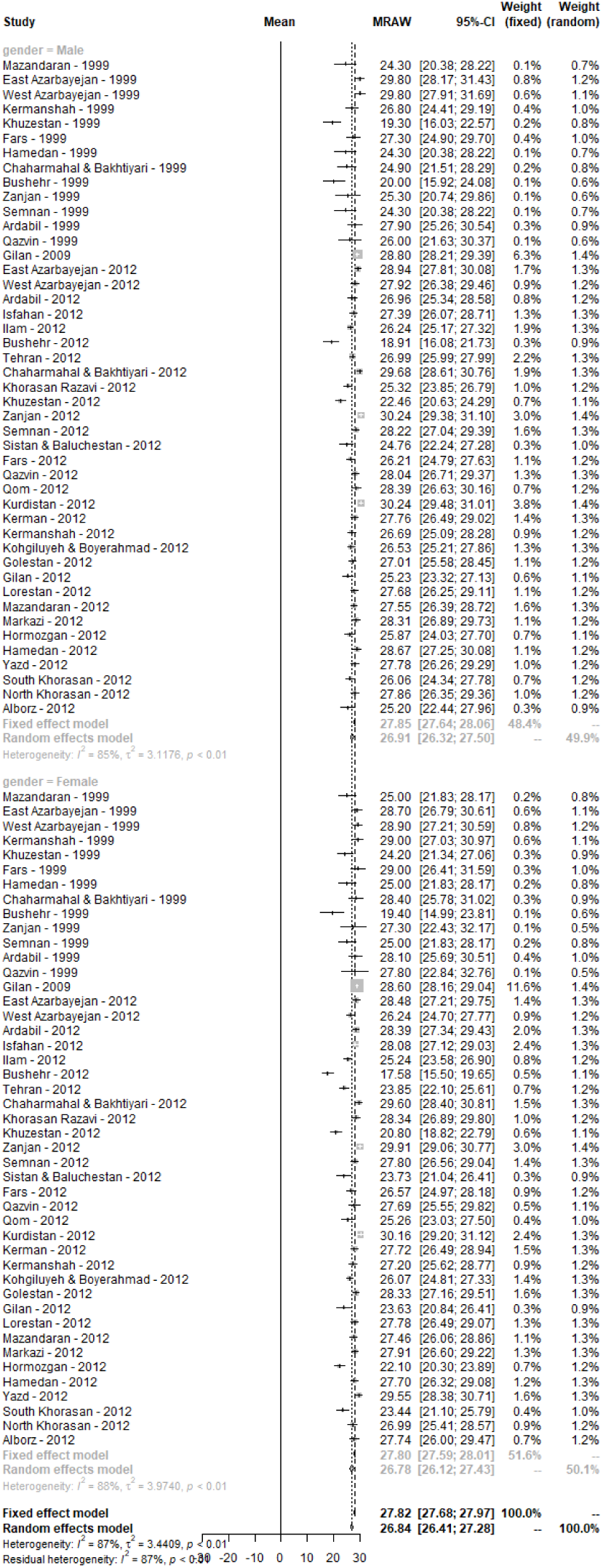
Forest plot for data reporting mean DMFT index of the elderly in all provinces of Iran.

**Figure 3.**
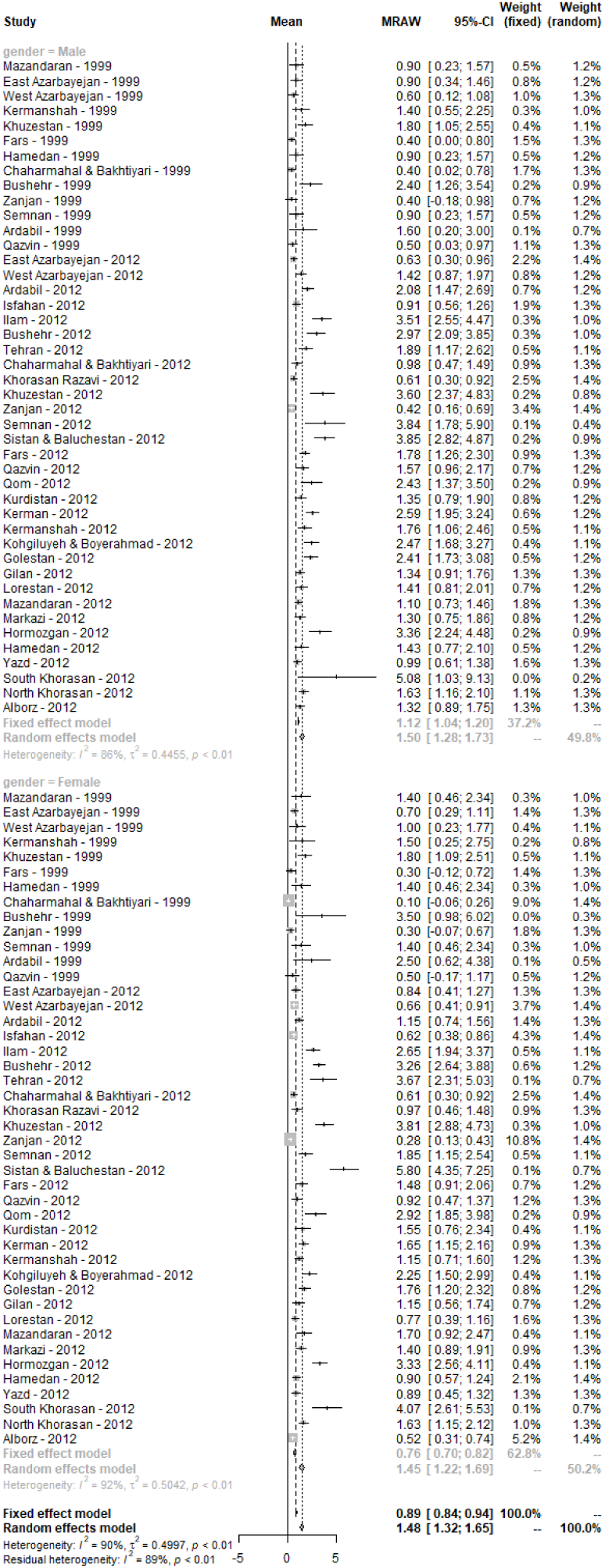
Forest plot for data reporting mean number of decayed teeth of the elderly in all provinces of Iran.

**Figure 4.**
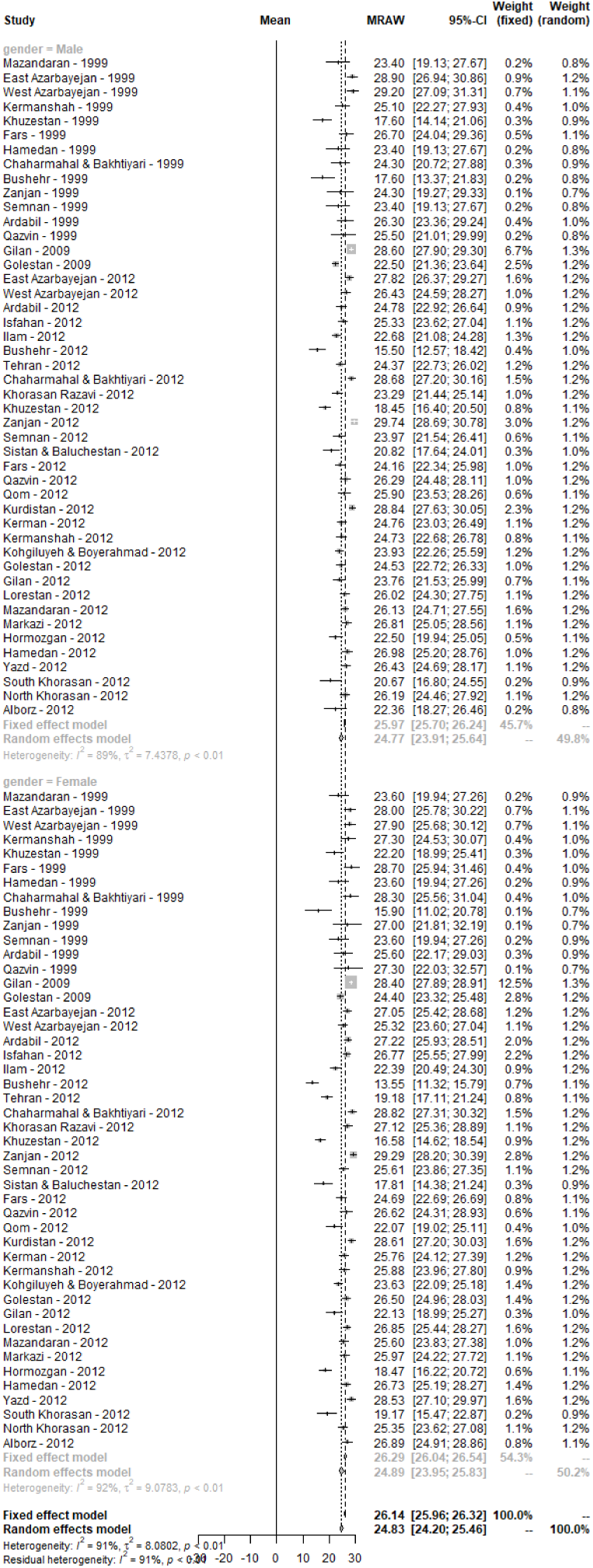
Forest plot for data reporting mean number of missing teeth of the elderly in all provinces of Iran.

**Figure 5.**
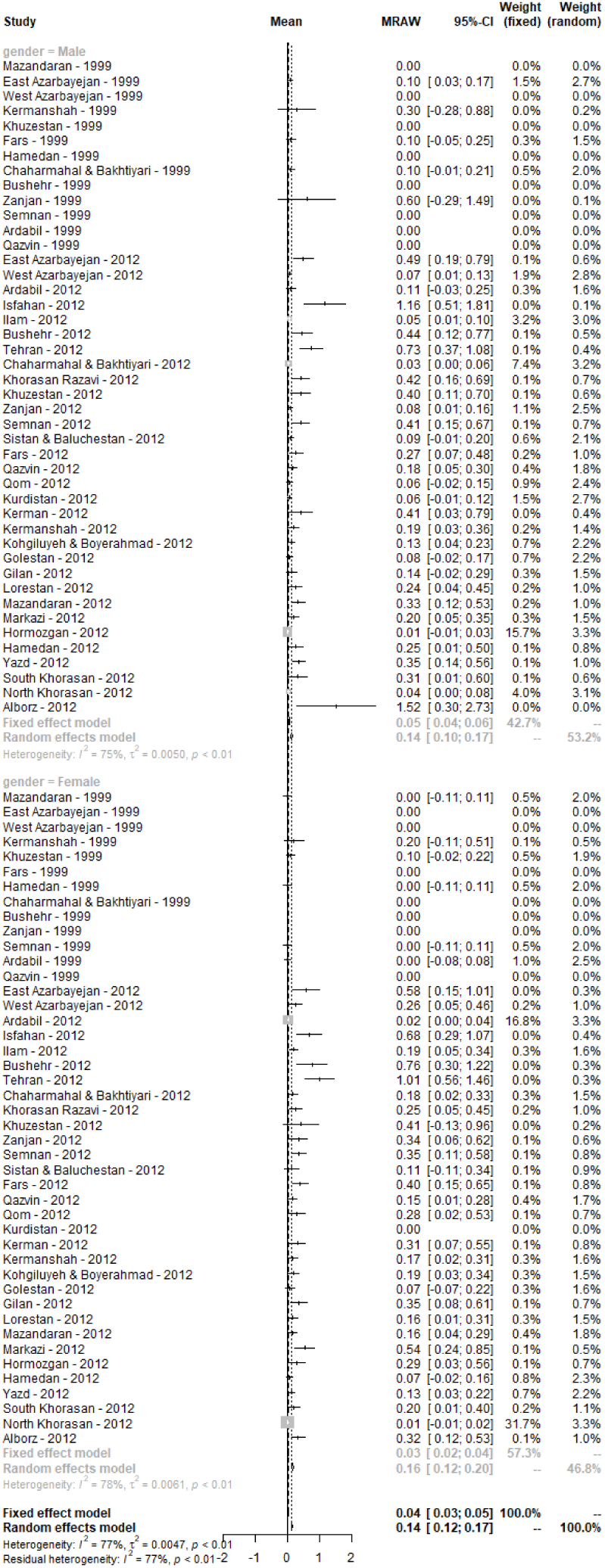
Forest plot for data reporting mean number of filled teeth of the elderly in all provinces of Iran.

The measured I^2^ for DMF was 87% which was statistically significant (P < 0.01). The map of mean DMF for each province is shown in Figure 6. The heterogeneity was also high for all components of DMF. Subgroup analysis was performed for detecting of resource of heterogeneity. For this reason, the population of Iran was divided into four regions of Southeast, North-Northeast, West, and Central (each region was homogeny in terms of socioeconomic variables based on years of schooling, employment rates, and family assets (21)). This regional analysis was heterogenous, either (Table 1 and Appendix 2).

**Table 1.**
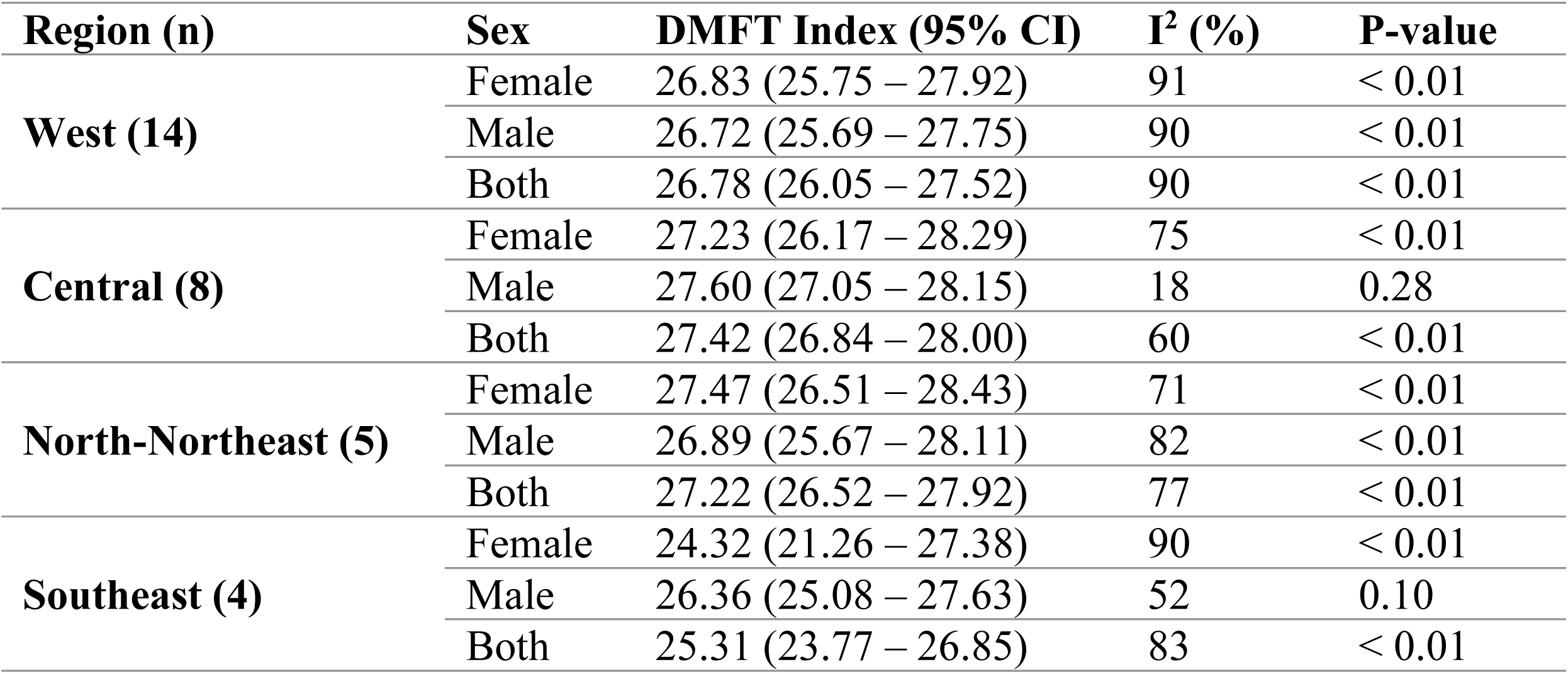
DMFT Index based on socioeconomic regions and sex

| **Region (n)** | **Sex** | **DMFT Index (95% CI)** | **I<sup>2</sup> (%)** | **P-value** |
| --- | --- | --- | --- | --- |
| **West (14)** | Female | 26.83 (25.75 – 27.92) | 91 | < 0.01 |
|  | Male | 26.72 (25.69 – 27.75) | 90 | < 0.01 |
|  | Both | 26.78 (26.05 – 27.52) | 90 | < 0.01 |
| **Central (8)** | Female | 27.23 (26.17 – 28.29) | 75 | < 0.01 |
|  | Male | 27.60 (27.05 – 28.15) | 18 | 0.28 |
|  | Both | 27.42 (26.84 – 28.00) | 60 | < 0.01 |
| **North-Northeast (5)** | Female | 27.47 (26.51 – 28.43) | 71 | < 0.01 |
|  | Male | 26.89 (25.67 – 28.11) | 82 | < 0.01 |
|  | Both | 27.22 (26.52 – 27.92) | 77 | < 0.01 |
| **Southeast (4)** | Female | 24.32 (21.26 – 27.38) | 90 | < 0.01 |
|  | Male | 26.36 (25.08 – 27.63) | 52 | 0.10 |
|  | Both | 25.31 (23.77 – 26.85) | 83 | < 0.01 |

**Figure 6.**
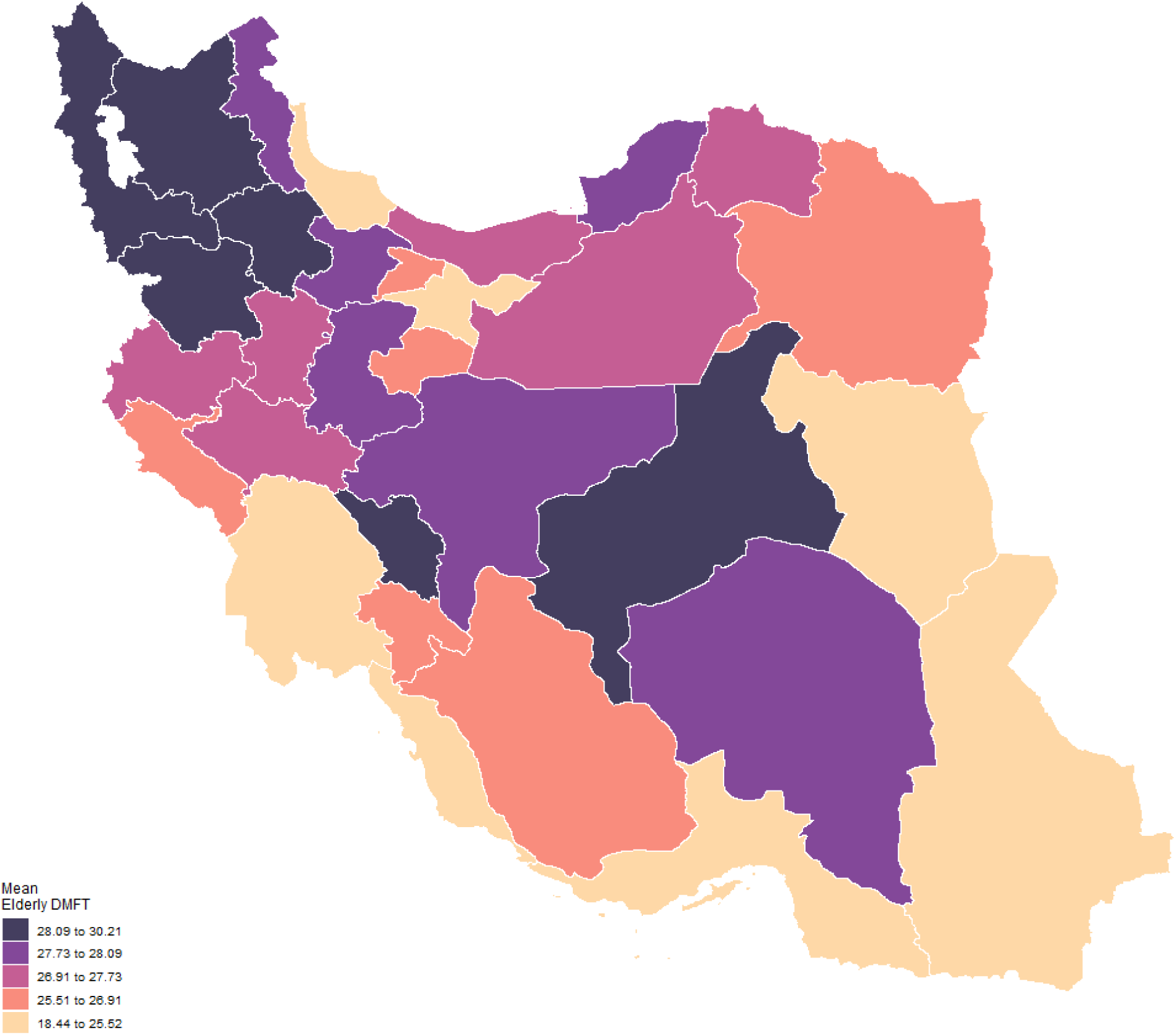
Map of mean DMFT index of the elderly in all provinces of Iran.

The results of Egger’s test were significant for DMF and its components which shows that there was considerable publication bias (Figure 7).

**Figure 7.**
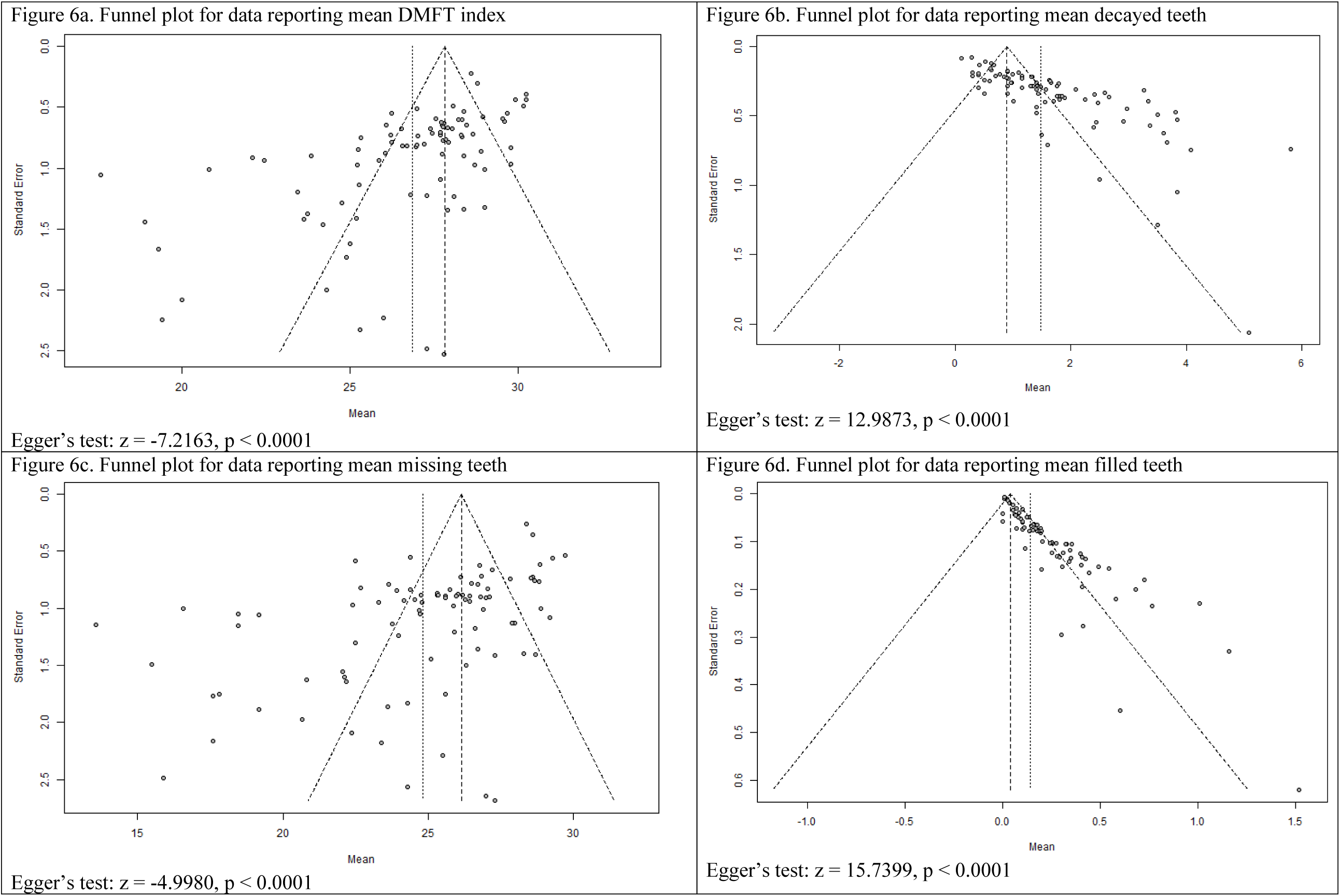
Funnel plots for data reporting mean DMFT index and its components of elderly in all provinces of Iran.

## Discussion

This pooled analysis is the first study reporting the DMFT index of Iranian elderly nationally and subnationally. We found a high mean DMFT index in the elderly Iranian population (26.84), which missing teeth accounted for 24.83 teeth (92.5% of DMFT). This means Iranian elderly on average have about seven teeth in their mouth, counting third molars.

We found little information regarding the DMFT index of elderly in the literature. Indeed, most studies we found had only reported the number of missing teeth. Hence, we used data of two extensive studies which were conducted nationwide in 1999 and 2012 and data of a study conducted in one province in 2009.

The calculated mean DMFT is high; however, studies from other countries have reported DMFT of their elderly population in a range near our results. For example, this index was 25.8 in Turkish elderly (22) and 25.4 and 27 in Norwegians and Croatians, respectively (23, 24). Some studies reported a higher or lower DMFT such as Brazil for 30.2 (25) and Australia for 18.9 (26). Similar to this study, missing teeth were responsible for the majority of DMFT in all these populations.

In developed countries, the portion of filled teeth is relatively higher than in developing countries (23, 27, 28) as they have more disciplined oral healthcare systems. The mean number of filled teeth in this study is 0.14, which accounts for 0.5% of the whole mean DMFT. This can show two things; first, there is limited access to operative dentistry services, and the other, that there still may be a belief in the Iranian community that they should extract carious teeth instead of filling it.

There was a significant disparity between provinces that Bushehr (18.44) had the lowest and Kurdistan (30.21) had the highest mean DMFT index. To explain this, we ran a secondary analysis based on the socioeconomic status of each region. This analysis did not address this difference, either. There is evidence that water fluoridation may prevent dental caries in children and adults (29). In a study measuring the amount of fluoride in groundwater resources in Iran, Bushehr had the highest level of fluoride and Kurdistan was among the provinces with the lowest amount of fluoride (30). This can be an explanation; however, further investigations are needed.

A relatively higher mean DMFT index was observed in males (26.91 compared to 26.78) which was not statistically significant. Men had slightly higher decayed teeth but slightly lower missing and filled teeth. Although there is no evident gender-based inequality, it should not be assumed to be a favourable matter since both genders have unfavourable mean DMFT index. In other words, both “fairness” and “goodness” are essential to health systems performance (31), and we do not see “goodness” in elderly dental caries in both genders.

Generally, DMFT increases with age. Mean DMFT index of 12- and 18-year-olds of Iran is reported to be 1.9 (32) and 4.3 (33), respectively. In these ages, decayed teeth are responsible for the majority of the DMFT index. DMFT is increasing to 11.0 in 35-to 44-year-olds which 60% it is due to missing teeth (34). Findings of our study show that elderly DMFT is over twofold of the reported adults DMFT. Besides, the number of decayed teeth decreases from 2.6 to 1.45 and the number of filled teeth is almost one-thirteenth (0.14 compared to 1.8). This means many sound, filled and decayed teeth will become hopeless and be extracted with ageing for different reasons including dental caries, periodontal diseases, inadequate functional ability of the remaining teeth, tendency to replace natural teeth with complete dentures because of cultural or economic factors, or little public or insurance support for oral health services.

### Limitation

Our analysis showed that there is a high publication bias in the data. The data were mainly from two national studies and there were limited data in the literature reporting DMFT of the elderly and its components. Another limitation was the quality of data, as studies have not reported their sampling and evaluation methods in details. Moreover, there is no definite assurance that the missing teeth component of the DMF index in the elderly is majorly due to dental caries or other factors such as periodontal diseases or dental trauma.

### Policy implications

The finding of high DMFT index in the elderly and vast disparity between provinces is of interest to policymakers. With the growing number of the elderly in the Iranian society which estimated to elevate from 6.1% in 2017 to 11% by 2036 (35), clearly, there should be more efforts to decrease the level of caries experience among the elderly. Dental caries are preventable so that preventive programmes should be encouraged first and then restorative services and then many teeth can be saved. Furthermore, targeted approaches may be attractive to policymakers since they can limit resource allocation and may be determined as addressing important determinants such as behaviour and access to dental care (with dental insurance coverage) (36).

Oral healthcare is mainly private in Iran especially in urban areas (80% private). In rural areas, 70% of oral healthcare, mostly basic oral services, are delivered by the public sector (37). Although it is estimated that nearly 90% of Iranian have social insurance coverage (38), insurance funds for oral healthcare are limited. However, there are few dental insurance plans which everyone including elderly can use them to cover their out of pocket expenditures, but it is not sufficient. We encourage oral health policymakers to advocate for oral health plan in a national or province-level considering the nature of oral disease which has large differences with other health conditions (39); for example, deductibles and copayments should be avoided and patients should be encouraged to use their oral health insurance regularly as delayed treatment can worsen their oral health status.

As the elderly population suffer from many other NCDs, it is an excellent practice to follow the common risk factor approach (40). This approach addresses risk factors that are in common with other chronic conditions. However, this factor had a little impact regarding promoting oral health, and there should be more radical actions such as “strike out independently of the other NCDs” (41). Either way, we encourage Iranian policymakers to use the latest evidence and experiences from other countries to reduce the amount of dental caries which can also be a measure of the socio-economic status of individuals.

## Conclusion

The results of this meta-analysis showed that the level of dental caries in Iranian over 65 years old is very high, and the majority is related to the missing teeth. The lack of specifically developed strategy for elderly oral health promotion in Iran, and the limited dental insurance coverage, just restricted to tooth extraction and full denture, for this age group, leads the society through edentulousness.

Therefore, the level of dental caries should be a priority in Iran, and oral health care investment should be devoted to the planning of rules, strategies and programs for elderly oral health. The integration of oral health into national general health programs may be effective to improve the oral health status and quality of life of this population group. Overall, more emphasis should be placed on oral health policies and principles of preventive oral care in Iran.

## Data Availability

Data will be available upon request.

## Appendices

**Appendix 1.**
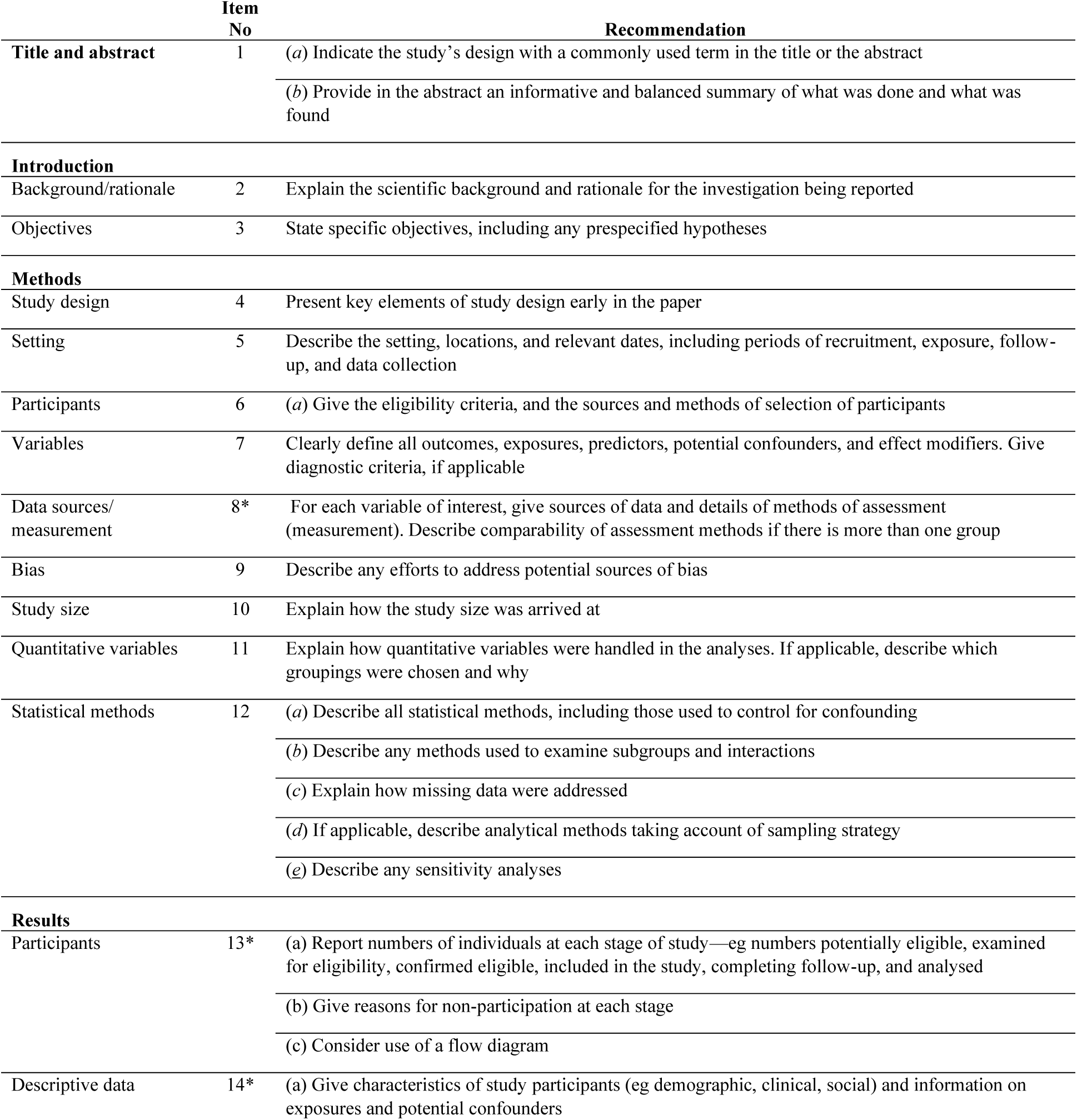
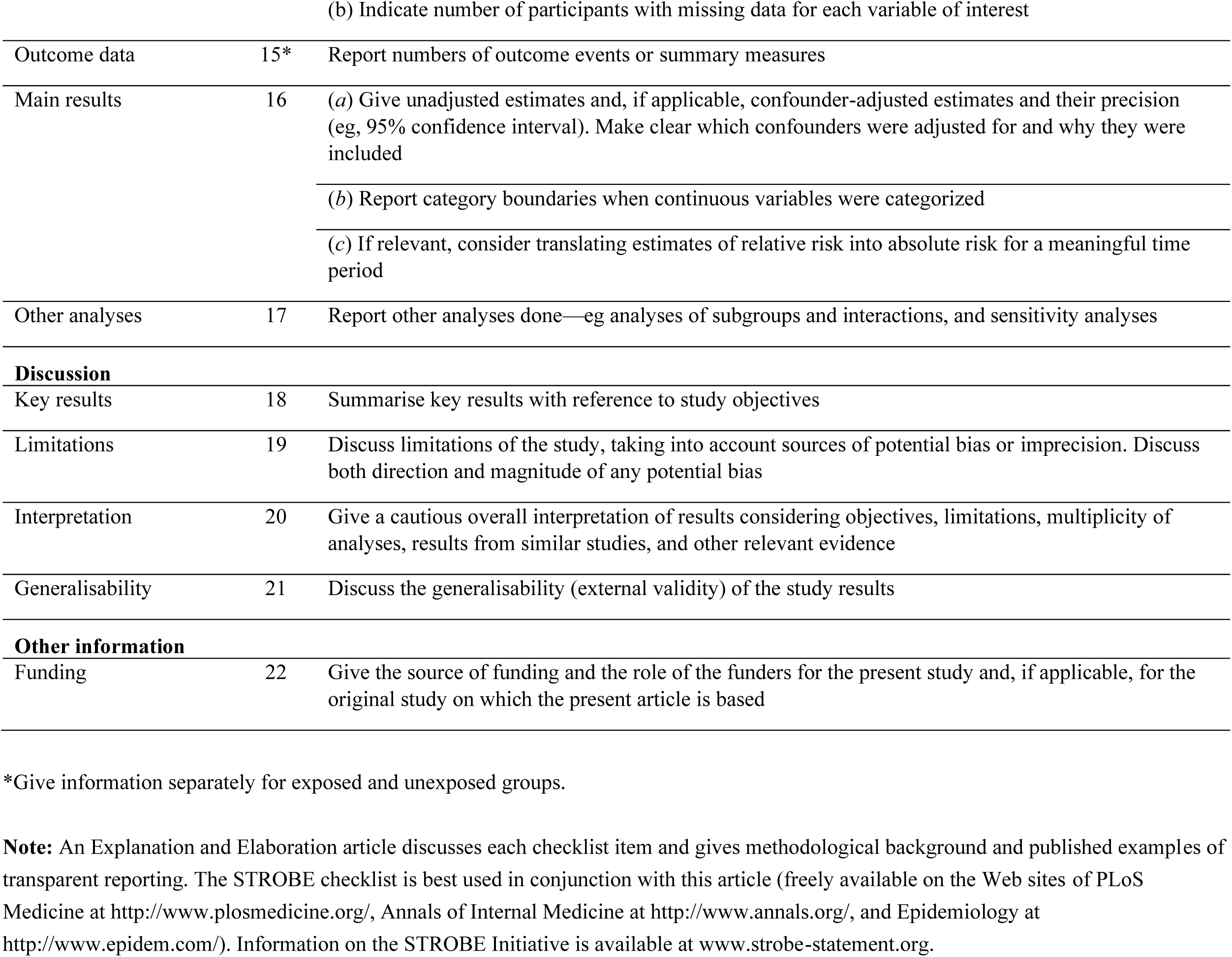
STROBE checklist. STROBE Statement—Checklist of items that should be included in reports of ***cross-sectional studies***

|  | Item No | Recommendation |
| --- | --- | --- |
| Title and abstract | 1 | (a) Indicate the study’s design with a commonly used term in the title or the abstract |
|  |  | (b) Provide in the abstract an informative and balanced summary of what was done and what was found |
| Introduction |  |  |
| Background/rationale | 2 | Explain the scientific background and rationale for the investigation being reported |
| Objectives | 3 | State specific objectives, including any prespecified hypotheses |
| Methods |  |  |
| Study design | 4 | Present key elements of study design early in the paper |
| Setting | 5 | Describe the setting, locations, and relevant dates, including periods of recruitment, exposure, follow-up, and data collection |
| Participants | 6 | (a) Give the eligibility criteria, and the sources and methods of selection of participants |
| Variables | 7 | Clearly define all outcomes, exposures, predictors, potential confounders, and effect modifiers. Give diagnostic criteria, if applicable |
| Data sources/measurement | 8* | For each variable of interest, give sources of data and details of methods of assessment (measurement). Describe comparability of assessment methods if there is more than one group |
| Bias | 9 | Describe any efforts to address potential sources of bias |
| Study size | 10 | Explain how the study size was arrived at |
| Quantitative variables | 11 | Explain how quantitative variables were handled in the analyses. If applicable, describe which groupings were chosen and why |
| Statistical methods | 12 | (a) Describe all statistical methods, including those used to control for confounding |
|  |  | (b) Describe any methods used to examine subgroups and interactions |
|  |  | (c) Explain how missing data were addressed |
|  |  | (d) If applicable, describe analytical methods taking account of sampling strategy |
|  |  | (e) Describe any sensitivity analyses |
| Results |  |  |
| Participants | 13* | (a) Report numbers of individuals at each stage of study—eg numbers potentially eligible, examined for eligibility, confirmed eligible, included in the study, completing follow-up, and analysed |
|  |  | (b) Give reasons for non-participation at each stage |
|  |  | (c) Consider use of a flow diagram |
| Descriptive data | 14* | (a) Give characteristics of study participants (eg demographic, clinical, social) and information on exposures and potential confounders |
| (b) Indicate number of participants with missing data for each variable of interest |  |  |
| Outcome data | 15* | Report numbers of outcome events or summary measures |
| Main results | 16 | (a) Give unadjusted estimates and, if applicable, confounder-adjusted estimates and their precision (eg, 95% confidence interval). Make clear which confounders were adjusted for and why they were included<br>(b) Report category boundaries when continuous variables were categorized<br>(c) If relevant, consider translating estimates of relative risk into absolute risk for a meaningful time period |
| Other analyses | 17 | Report other analyses done—eg analyses of subgroups and interactions, and sensitivity analyses |
| **Discussion** |  |  |
| Key results | 18 | Summarise key results with reference to study objectives |
| Limitations | 19 | Discuss limitations of the study, taking into account sources of potential bias or imprecision. Discuss both direction and magnitude of any potential bias |
| Interpretation | 20 | Give a cautious overall interpretation of results considering objectives, limitations, multiplicity of analyses, results from similar studies, and other relevant evidence |
| Generalisability | 21 | Discuss the generalisability (external validity) of the study results |
| **Other information** |  |  |
| Funding | 22 | Give the source of funding and the role of the funders for the present study and, if applicable, for the original study on which the present article is based |

\*Give information separately for exposed and unexposed groups.
**Note:** An Explanation and Elaboration article discusses each checklist item and gives methodological background and published examples of transparent reporting. The STROBE checklist is best used in conjunction with this article (freely available on the Web sites of PLoS Medicine at http://www.plosmedicine.org/, Annals of Internal Medicine at http://www.annals.org/, and Epidemiology at http://www.epidem.com/). Information on the STROBE Initiative is available at www.strobe-statement.org.

**Appendix 2A.**
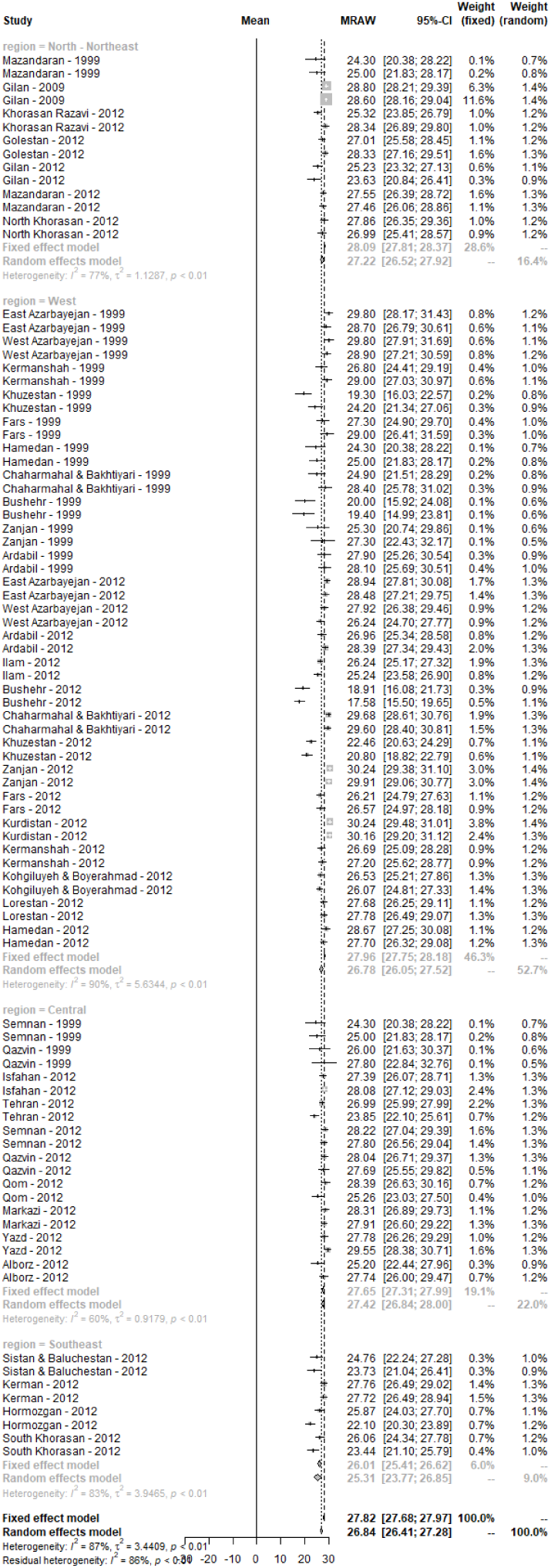
DMFT Index based on socioeconomic regions and sex (forest plot) – Both genders

**Appendix 2B.**
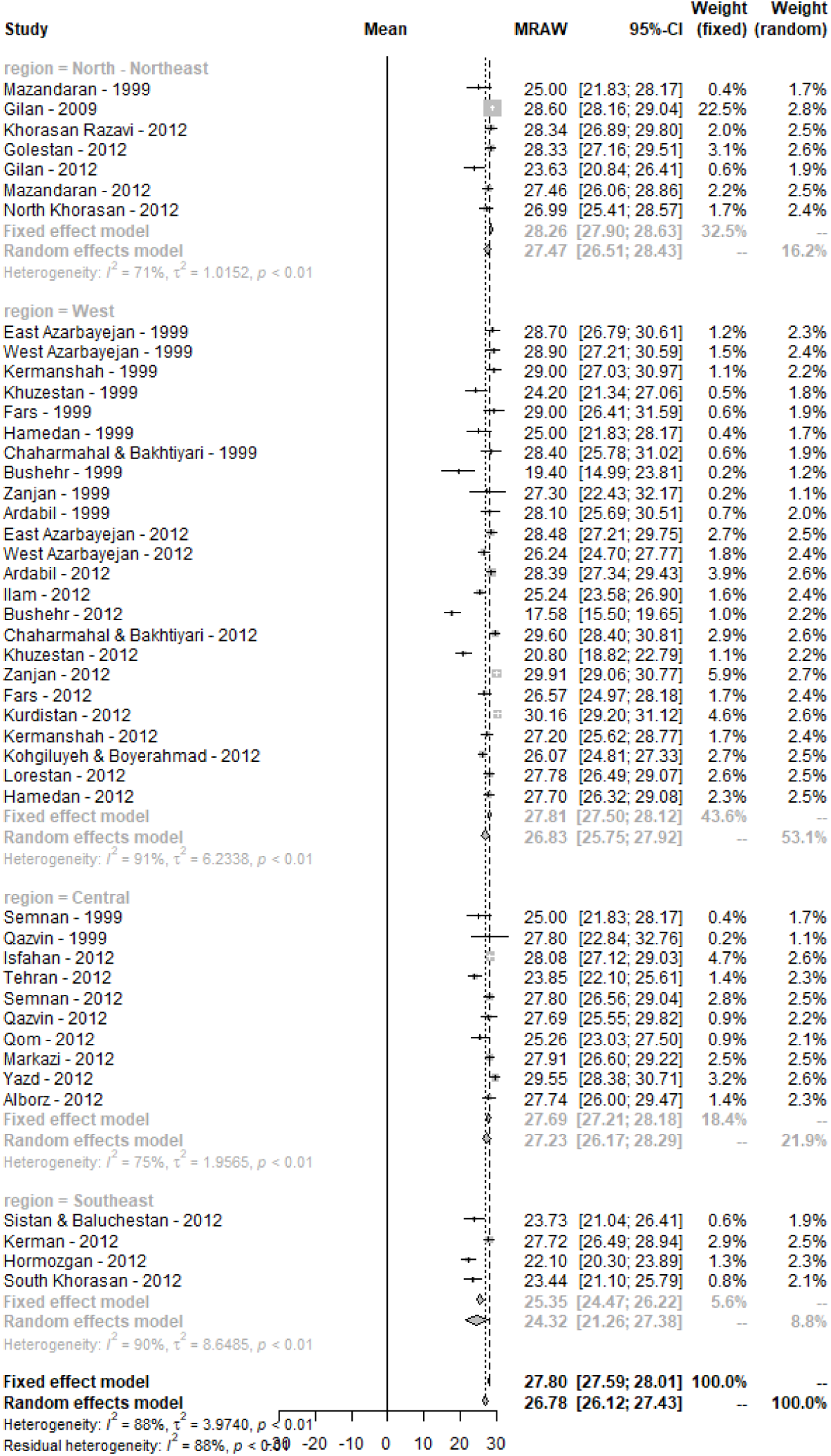
DMFT Index based on socioeconomic regions and sex (forest plot) – Females

**Appendix 2C.**
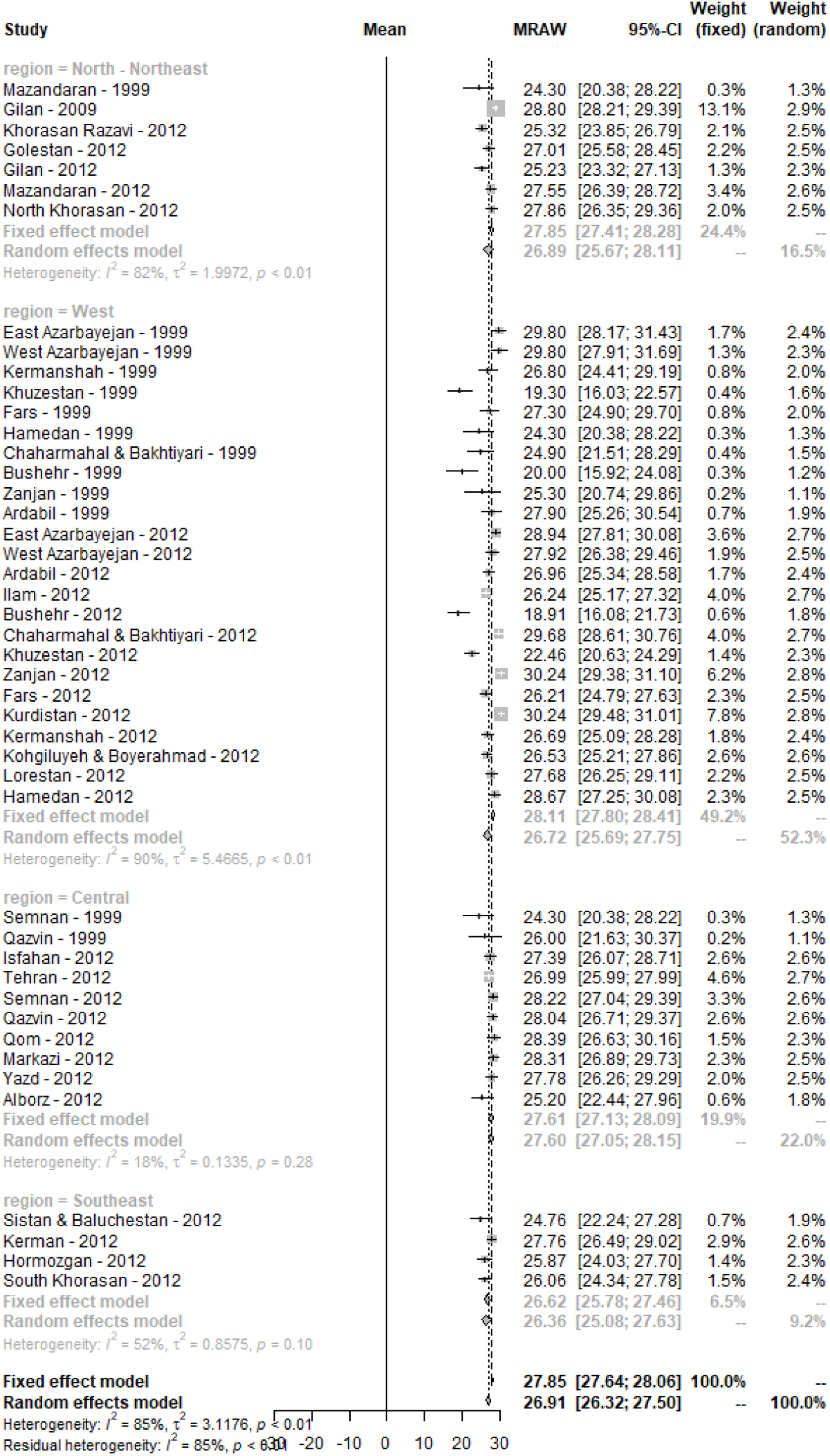
DMFT Index based on socioeconomic regions and sex (forest plot) – Males

